# Scoping national research infrastructure to inform the design and delivery strategy for a platform clinical trial in Parkinson’s disease

**DOI:** 10.64898/2026.06.04.26354792

**Authors:** Rebecca Petty, Marie-Louise Zeissler, Veena Agarwal, Jennifer Allison, Sandra Bartolomeu-Pires, Michèle Bartlett, Rebecca Croucher, Helen Collins, Sally Collins, Emma Davies, Joy Duffen, Romy Ellis-Doyle, Cristina Gonzalez-Robles, Jemma Inches, Laurel Miller, Georgia Mills, Sheila Wonnacott, Thomas Foltynie, Camille B. Carroll, Stephen Mullin, the EJS ACT-PD Consortium

## Abstract

**Objective:** To map national Parkinson’s disease (PD) research capability to inform an inclusive delivery strategy for a large-scale clinical trial.

**Background:** Few people with PD participate in clinical trials, particularly from under-served populations. The Edmond J Safra Accelerating Clinical Trials in PD initiative (EJS ACT-PD) aims to deliver an inclusive multi-arm multi-stage (MAMS) disease modification PD trial.

**Methods:** A survey disseminated to National Health Service (NHS) hospitals assessed PD research capability regarding trial experience, rater expertise, trial facilities and specialist investigations. A process was developed to categorise sites into 3 tiers, with tier 1 having the least PD-research capability or experience, and tier 3 being experienced specialist centres. We mapped tiers to PD prevalence, social deprivation and ethnic diversity to identify infrastructure gaps. We developed trial delivery strategies to facilitate rapid and inclusive recruitment.

**Results:** Out of 97 survey responses, 43 sites were categorised as tier 1, 33 as tier 2 and 21 as tier 3. Diversity and social deprivation index were higher for tier 3 sites (predominantly urban). A greater proportion of tier 1 and 2 sites were situated in areas of higher PD prevalence (predominantly rural). Ninety one percent of sites reported experience with remote trial delivery methods. Our delivery strategy included: initial trial set-up at tier 3 sites to enable rapid and ethnically diverse recruitment; core funded staff within strategic sites to develop regional solutions for inclusive trial participation and to enable research opportunity provision in areas where currently very little exists, and a hybrid delivery model of in-person and remote study visits, ensuring maximal acceptability and deliverability.

**Conclusions:** The mapping of current PD research delivery capability has allowed us to develop a trial delivery strategy that will broaden the provision of research participation opportunity to under-served groups. It has also enabled existing infrastructure to be maximised while mitigating identified gaps.

## BACKGROUND

Clinical trials are often not inclusive, meaning that study populations are not representative; this limits the generalisability of findings[1] and risks perpetuating healthcare inequalities[2]. Barriers to inclusion exist at patient, trial protocol, and site delivery infrastructure level which can result in geographical exclusion[3-5]. Broadening access to trials requires an understanding of the trial delivery landscape beyond the most experienced sites.

Parkinson’s disease (PD) is a rapidly growing neurological condition[6]. This debilitating, slowly progressive disease causes multiple motor and non-motor symptoms that significantly impact people with PD and their families[7]. Despite considerable research efforts, no disease modifying therapy to stop or reverse progression has been identified. There is an urgent need to improve the efficiency with which disease modifying therapy trials are conducted. The Edmond J Safra Accelerating Clinical Trials in Parkinson’s Disease (EJS ACT-PD) consortium[8] developed a platform trial to evaluate disease modifying therapies in PD. The EJS ACT-PD consortium consisted of 90 members across six working groups, including representation from clinicians with an interest in PD research, funders, trial delivery staff, PD charities and those with lived experience of PD. The trial aims to recruit 1600 participants across four treatment arms. The ambition is to inclusively recruit a representative population of participants from across the United Kingdom (UK), in particular with regards to age, sex, social deprivation, ethnicity and geographical location. A detailed understanding of infrastructure, barriers and facilitators to research delivery within the UK is required to inform trial design decisions and delivery strategies.

Here we investigated site-level capability for PD research delivery at infrastructure level in terms of skills, experience and resources that might make research opportunity less inclusive from a geographical perspective, and how this intersected with population-level ethnicity, deprivation and disease prevalence. These findings are used to inform trial design decisions and delivery strategy to ensure trial findings are generalisable.

## METHODS

An infrastructure working group (IFWG) was convened within the consortium, consisting of multi-professional members with broad geographical representation including clinicians with an interest PD research, National Institute for Health and Care Research (NIHR) representatives, trial delivery nurses and allied health professionals, clinicians and trial managers, PD charities as well as patient and lived experience experts. The group’s remit included exploring requirements to ensure the trial’s recruitment and delivery objectives could be met.

### Survey Development

The IFWG developed a survey, informed by two key resources: 1) a systematic scoping review of clinical trial methodology for PD disease modification trials in PD[9] and 2) NIHR Clinical Research Network (CRN) guidelines for site expression of interest surveys (a UK standard process for the assessment and rapid identification of suitable trial delivery sites)[10]. Questions were also added or adapted in response to lived experienced experts’ feedback. This resulted in the identification of domains pertaining to PD trial experience (overall PD research experience including past recruitment and staffing; rater experience in using PD-specific and generic outcome measures; remote trial delivery methods; digital outcome measures; intravenous and intrathecal drug delivery; cerebral spinal fluid (CSF) collections) and facilities (imaging equipment; sample/processing storage; trial accommodation and research space; trial pharmacy access).

A web-based survey was created on REDCap (Research Electronic Data Capture) online surveys; all questions were mandatory to reduce missing data. The survey was pre-tested by the IFWG members and refined prior to dissemination to ensure questions were understandable and easy to complete.

### Survey Dissemination

The survey was primarily disseminated to hospitals, research active organisations and research teams via national clinical research networks in England, Scotland, Wales and Northern Ireland. National research network staff were asked to encourage completion of surveys within their regions and provide information on which organisations were active in neurology research. Organisations with previous experience of delivering phase 2 and 3 drug trials of disease modifying therapies in PD over the past five years were additionally identified from a national dataset and prioritised for further contact via phone by the EJS ACT-PD team and email if they had not responded to the survey. In a second round of dissemination to encourage organisations who had not responded, the survey was sent to personal contacts compiled from the EJS ACT-PD consortium and through the Association of British Neurologists Movement Disorders Special Interest Group.

### Data Analysis

#### Organisation characteristics

Organisations were classified into 13 types using a UK classification system as per the NIHR Reporting Platform National Health Service (NHS) Trust classifications[11]. An organisation was considered active in PD research if it had delivered at least one phase 2 or 3 PD Clinical Trial of an Investigational Medicinal Product in the five years prior to 25th November 2022. The percentage of organisations within each category and overall percentage of PD research active organisations which returned a survey response was then determined.

#### Survey response data

As an NHS organisation could consist of multiple individual hospital sites, survey response rate was measured by site rather than organisation. Survey data were presented as absolute numbers and percentages of site level survey responses (unless otherwise stated) for binary and categorical variables). A data dictionary and coding definitions for quantitative data fields can be found in additional file 1. For some questions respondents were able to provide optional free-text responses, which were thematically analysed.

#### Tier categorisation

Sites were categorised into 3 tiers based on their capability to deliver PD trials. Tier 3 sites were specialist neuroscience centres with extensive PD trial delivery experience; tier 2 sites had experience of conducting PD trials but may have lacked specialist personnel or equipment; and tier 1 sites had limited capability and no previous experience of PD trial delivery. Details of specific survey questions and answer options for categorisation into the tiers is available in additional file 1.

#### Mapping prevalence of under-served groups

Publicly available datasets were used to map prevalence of groups that often lack opportunity to be involved in research. This included those living in rural locations, with low income, or from minority backgrounds[12]. Ethnicity data were extracted from the Office for National Statistics 2021 Census Ethnic Group Table[13] and presented as the proportion of non-white individuals by Local Authorities District. PD prevalence was sourced from the UK Parkinson’s Excellence Network data dashboards[14] and presented as the percentage of the population with PD by English Clinical Commissioning Group region and the devolved nations. Social deprivation was described using the Index of Multiple Deprivation (IMD). The IMD is a multicomponent score that ranks areas from least to most deprived areas. Postcode checkers for England[15], Scotland[16], Wales[17] and Northern Ireland[18] were used to identify a site’s IMD decile, with a lower decile indicating a greater level of social deprivation.

### Development of mitigation strategies

The findings of the survey were used to inform the development of strategies to mitigate identified challenges to rapid and inclusive trial recruitment. Strategies were presented to and discussed by the consortium steering group, who then recommended their adoption in the trial.

### Patient and Public Involvement

People with PD and family members were involved as advisors within the EJS ACT-PD project as part of the consortium’s Patient and Public Involvement and Engagement (PPIE) Working Group as well as within the IFWG. The project implemented a detailed PPIE evaluation strategy[19] and allowed for their involvement in the formulation of trial delivery recommendations for the EJS ACT-PD platform trial that have resulted from this work.

## RESULTS

### Response Rate

Survey responses were received from 110/249 (44.2%) NHS organisations (112 individual site responses). Nine of the individual site responses were incomplete with sites indicating that they did not have the capability to carry out neurological research and were not interested in expanding into this area.

A further six sites indicated they were not willing to complete the survey at all for the following reasons: not having staff available to complete the survey (n=2), not having any Principal Investigator (PI) oversight available (n=1), no capability to run studies involving an investigational medicinal product (IMP) (n=1) and no reason given (n=2). This resulted in 97 site level surveys from 95 NHS organisations being fully completed.

### Organisation Characteristics

The types of organisation that completed the survey are summarised in Table 1. Most responses were received from medium acute (n=25/97 (26.3%)) and large acute (n=20/97 (20.6%)) organisations. Seventy-five organisations were identified from the NIHR Reporting Platform as having been PD research active within the past five years, 57/75 (76.0%) of which returned a survey response. Responses were received from a further 38 organisations not listed on the database.

**Table 1:** Number of completed surveys per Trust type, out of all 97 site level responses.

| Organisation Type<sup>a</sup> | Site level responses |  |  | Total number of national organisations<sup>b</sup> |
| --- | --- | --- | --- | --- |
|  | N | % of all 97 site responses | % site level responses per number of national organisations within each organisation type |  |
| Large Acute | 20 | 20.6 | 43.5 | 46 |
| Medium Acute | 25 | 26.3 | 48.1 | 52 |
| Small Acute | 8 | 8.2 | 28.6 | 28 |
| Acute teaching | 13 | 13.4 | 50.0 | 26 |
| Welsh Health Board | 6 | 6.2 | 85.7 | 7 |
| Scottish Regional Health Board | 5 | 5.2 | 33.3 | 15 |
| Northern Irish NHS Trust | 1 | 1.0 | 16.7 | 6 |
| Specialist Trust<sup>c</sup> | 17 | 17.5 | 47.7 | 111 |
| Other<sup>d</sup> | 2 | 2.0 | 60.0 | 12 |

<sup>a</sup>Trust categories extracted from the NIHR Reporting Platform. Categorisation of trust type is inconsistent and therefore no current definition exists
<sup>b</sup>Data extracted from the NIHR Reporting Platform on 10.10.2025
<sup>c</sup>Includes mental health, care, ambulance and acute specialist including paediatric specialty
<sup>d</sup>Includes acute and 'other' trusts

### Experience and Facilities

Findings are summarised in additional file 2.

#### Trial experience

Eighty four out of 97 (86.6%) sites (31/43 (72.1%) tier 1 sites, 32/33 (97.0%) tier 2 sites and 21/21 (100%) tier 3 sites) reported capability to deliver neurological research. Almost all sites (89/97 site responses (91.8%), 38/43 (88.4%) tier 1 sites, 30/33 (90.9%) tier 2 sites and 21/21 (100%) tier 3 sites) had experience of delivering remote methods in trials.

#### Rater experience

Rater capacity was 44/97 (45.4%) across all sites with (10/43 (23.3%) tier 1 sites, 19/33 (57.6%) tier 2 sites, 15/21 (71.4%) tier 3 sites) having sufficient staff to provide a blinded rater and a backup rater. Overall, slightly more sites were experienced with generic measures (a median of 68, IQR 13 sites) compared to with PD specific measures (a median of 61, IQR 13 sites). Analysis of 46 free-text responses related to workforce challenges revealed a need for additional training (n=11/46 sites (23.9%)), lack of experience (n=11/46 (23.9%)), lack of staff (n=7/46 (15.2%)), and issues related to concurrent research workload (n=5/46 (10.9%)).

Roughly half of the sites had experience with wearables (n=46/97 (47.4%)) and internet apps (n=48/97 (49.5%)), with slightly more having experience with smartphone apps (n=59/97 (60.8%)). Two sites (2/97 (2.1%)) did not have Wi-Fi in the area used to deliver research. Thirty-two sites provided free-text responses relating to use of digital measures. Themes included: difficulty connecting to Wi-Fi or uploading data (13/32 (40.6%)), websites or applications blocked due to NHS firewalls or security issues (8/32 (25%)); software issues (8/32 (25%)) and hardware issues (3/32 (9.4%)).

#### Trial Facilities

All tier 2 and 3 sites and nearly all tier 1 sites (34/41 (90.7%)) had access to a clinical trial pharmacy, while just over half had dedicated research space (47/97 (58.8%)), (17/43 (39.5%) tier 1 sites, 21/33 (63.6%) tier 2 sites and 19/21 (90.5%) tier 3 sites). Overnight accommodation was much less frequently available (15/97 site responses (15.5%), 4/43 (9.3%) tier 1 sites, 4/33 (12.1%) tier 2 sites and 7/21 (33.3%) tier 3 sites, particularly for care partners (10/97 (10.3%) sites, 4/43 (9.3%) tier 1 sites, 3/33 (9.1%) tier 2 sites and 3/21 (14.3%) tier 3 sites).

Fifty-six of the 97 sites (57.8%) were able to undertake home visits for research purposes. Themes related to home visits from free text responses (41 respondents) included barriers of lack of staff (n=22/41 (53.7%)) and NHS policy or insurance issues (n=7/41 (17.1%)).

### Specialist Investigations

Tier 1 and 2 sites had less experience and capability in using Magnetic Resonance Imaging (MRI) and Dopamine active Transfer Scan (DaTSCAN) equipment, as well as undertaking Cerebral Spinal Fluid (CSF) collections (Table 2). However, in a separate open-ended question roughly 20% of all 97 sites stated they may be able to outsource scanning capacity within nearby acute Trusts or the private sector. Consistent with the criteria to be categorised a tier 3 site, all tier 3 sites had the experience and capability to undertake MRI and CSF investigations.

**Table 2:** Experience and capability of specialist investigations overall and by tiers.

|  | **Overall**<br>**n=97 (%)** | **Tier 1**<br>**n=43 (%)** | **Tier 2**<br>**n=33 (%)** | **Tier 3**<br>**n=21 (%)** |
| --- | --- | --- | --- | --- |
| **MRI** | 53(54.6) | 16(37.2) | 16(48.5) | 21(100) |
| **DaTSCAN** | 29(29.9) | 7(16.3) | 8(24.2) | 14(66.7) |
| **CSF** | 32(33.0) | 7(16.3) | 4(12.1) | 21(100) |

Abbreviations: MRI, Magnetic Resonance Imaging; CSF, Cerebrospinal Fluid; DaTSCAN, Dopamine Active Transfer Scan

### Mapping Sites to Ethnicity, PD Prevalence and Social Deprivation

There was representation of all site tier levels across England, Scotland and Wales, apart from one region in England with no tier 3 site. Data were returned from one site in Northern Ireland which was categorised as tier 1. Site locations were mapped relative to indices of PD prevalence, ethnicity and social deprivation (Table 3, Figures 1a and 1b).

**Table 3:** Number of tiered sites mapped to ethnicity, PD prevalence and social deprivation.

| Ethnicity (non-white) |  |  |  |  |
| --- | --- | --- | --- | --- |
| % of non-white | Total sites (n=97)<br>n(% of all sites) | Tier 1 (n=43)<br>n(% within each tier) | Tier 2 (n=33)<br>n(% within each tier) | Tier 3 (n=21)<br>n(% within each tier) |
| ≤10% | 42 (43.3) | 21 (48.8) | 16 (48.5) | 5 (19.0) |
| 10.1 – 20% | 22 (22.7) | 11 (25.6) | 6 (18.2) | 5 (28.6) |
| 20.1 – 30% | 14 (14.4) | 4 (9.3) | 6 (18.2) | 4 (19.0) |
| 30.1 – 40% | 8 (8.2) | 3 (7.0) | 2 (6.1) | 3 (14.2) |
| >40% | 5 (5.2) | 1 (2.3) | 2 (6.1) | 2 (9.5) |
| Unknown<sup>a</sup> | 6 (6.2) | 3(7.0) | 1(3.0) | 2(9.5) |
| PD Prevalence |  |  |  |  |
| % of population with PD | Total sites (n=97)<br>n(% of all sites) | Tier 1 (n=43)<br>n(% within each tier) | Tier 2 (n=33)<br>n(% within each tier) | Tier 3 (n=21)<br>n(% within each tier) |
| ≤0.20% | 36 (37.1) | 12 (27.9) | 10 (30.3) | 14 (66.7) |
| 0.21 – 0.30% | 55 (56.7) | 27 (62.8) | 21 (63.6) | 7 (33.3) |
| 0.31 – 0.40% | 5 (5.2) | 4 (9.3) | 1 (3.0) | 0 (0.0) |
| >0.40% | 1 (1.0) | 0 (0.0) | 1 (3.0) | 0 (0.0) |
| Social Deprivation |  |  |  |  |
| IMD Decile | Total sites (n=97)<br>n(% of all sites) | Tier 1 (n=43)<br>n(% within each tier) | Tier 2 (n=33)<br>n(% within each tier) | Tier 3 (n=21)<br>n(% within each tier) |
| 1-2 | 12 (12.4) | 6 (14.0) | 2 (6.0) | 4 (19.0) |
| 3-4 | 19 (19.6) | 9 (20.9) | 5 (15.2) | 5 (23.8) |
| 5-6 | 27 (27.8) | 10 (23.3) | 12 (36.4) | 5 (23.8) |
| 7-8 | 19 (19.6) | 8 (18.6) | 8 (24.2) | 3 (14.3) |
| 9-10 | 19 (19.6) | 9 (20.9) | 6 (18.2) | 4 (19.0) |
| Unknown<sup>b</sup> | 1 (1.0) | 1 (1.0) | 0 (0.0) | 0 (0.0) |

<sup>a</sup>Data unavailable for Scotland and Northern Ireland
<sup>b</sup>IMD decile unavailable for Northern Ireland

**Figure 1:**
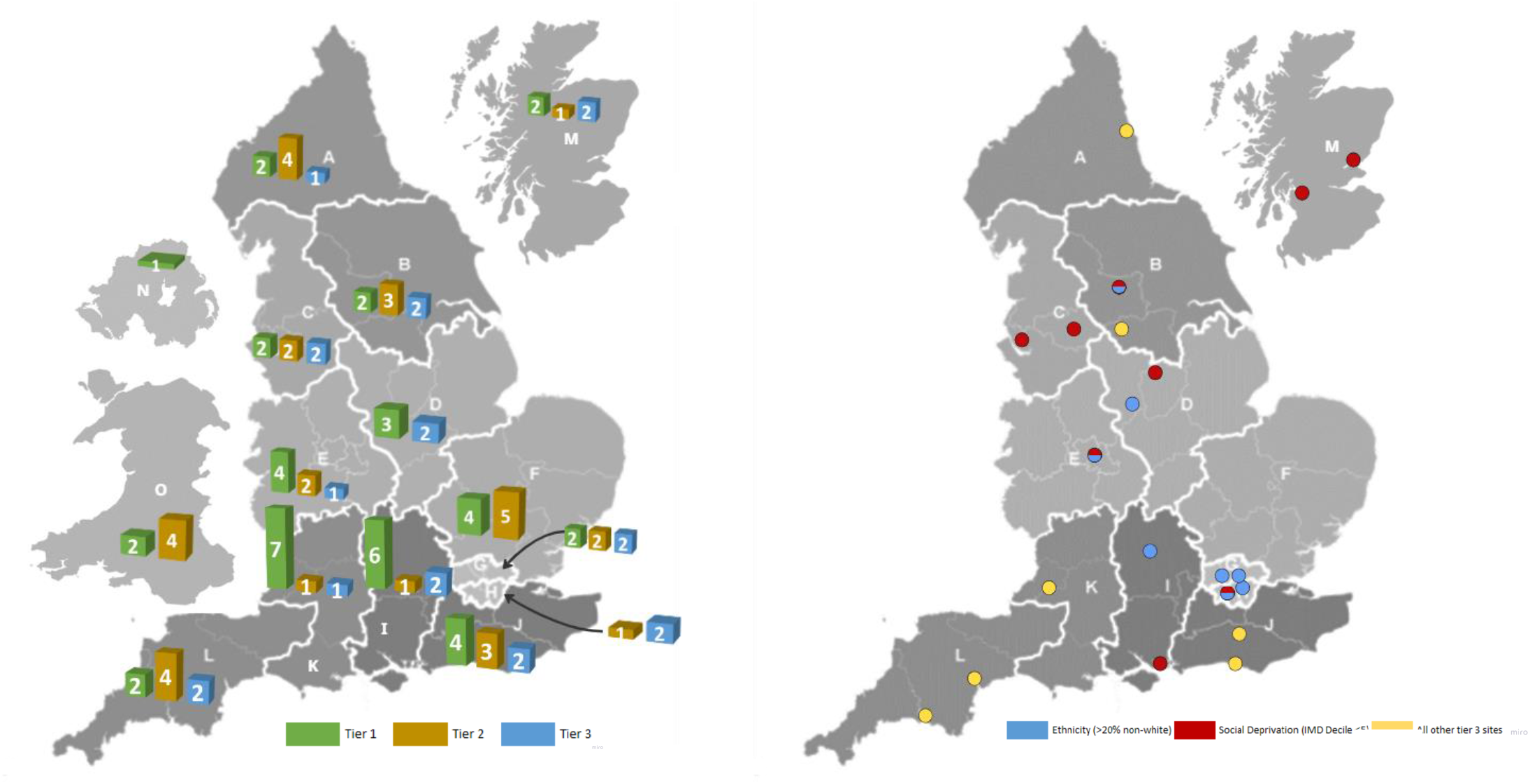
Distribution of tiered sites per RDN region and the devolved nations Figure 1b: Mapping of tier 3 sites to ethnicity and social deprivation. England Regions: A - North East and North Cumbria; B - Yorkshire & Humber; C - North West; D - East Midlands; E - West Midlands; F - East of England; G - North London; H - South London; I - South East Central; J - South East Coast; K - South West Central; L - South West Peninsula. Devolved Nations: M - Scotland; N - Northern Ireland; O – Wales

Overall, the majority of sites were situated in areas where the population of non-white was ≤10%. Of all tiers, tier 3 had the highest proportion of sites located in areas with a higher than 20% non-white population: 9/21(42.9%) tier 3 sites, 10/33(30.3%) Tier 2 and 8/43(18.6%) tier 1 sites. The overall number of sites in these areas was similar for all tiers (n=8, n=10 and n=9 for tier 1, 2 and 3 respectively).

In terms of social deprivation, the majority of sites across all tiers were situated in areas with average levels of deprivation (5th and 6th IMD decile, n=27)). A higher proportion of sites in lower IMD deciles (more deprived) (1-4) were tier 3 (n=9/21 (43%)).

Most tier 3 sites were located in areas with lower PD prevalence of less than 0.20% (n=14/21 (66.7% of tier 3 sites).

### Trial Strategies Implemented

Based on findings, challenges and opportunities for trial delivery were identified, and recommendations to mitigate the challenges and benefit from the opportunities were made by the IFWG (additional file 2) for implementation in the EJS ACT-PD trial:

#### Site selection strategy

It was determined that there would be at least one tier 3 site in each Research Delivery Natwork (RDN) region and devolved nation, thereby ensuring geographical distribution of PD trial delivery expertise and experience. Specialist biomarker and imaging sub-studies would be undertaken in selected tier 3 sites.

#### Core staff placement

Core funded staff would be located at strategic sites within each region, predominantly tier 3, able to upskill and mentor staff within less experienced sites. Additionally, these staff would support the development of recruitment and retention strategies for under-served groups within their regions, such as those from socially deprived and ethnic minority groups, or those living in coastal and rural communities. Development of a core staff community would allow for knowledge exchange and sharing of best practice.

#### A hybrid trial delivery model

Study visits would be delivered remotely or in-person (according to participant preference) in order to allow for maximum research opportunity. This would be particularly important for those in more geographically remote areas, or those with mobility restrictions/travel issues. The hybrid delivery model would be supported by:

- A central pharmacy to dispense trial medication directly to participants’ homes
- No OFF state assessments, to support participants who wish to undertake in-person study visits, and eliminate the possible need for overnight hospital accommodation

A study participant feedback questionnaire would be implemented to enable understanding of how the trial protocol decisions have impacted participant retention and satisfaction of trial participation.

## DISCUSSION

In this study we have mapped national PD research capability to inform the development of a delivery strategy that maximises inclusive access to a large-scale clinical trial. Mapping research capability to prevalence of under-served communities will assist with resource planning and help sites develop specific recruitment strategies and goals.

The majority of the survey respondents were tier 1 sites, predominantly situated outside of large urban conurbations, in locations with an older population[20]. All but one tier 1 site indicated interest in delivering future PD trials, highlighting untapped potential and enthusiasm to increase PD research delivery capacity across the UK in locations where PD is more prevalent. Individuals living in remote rural and coastal regions are currently under-served by health research, with poorer health outcomes and lower engagement with healthcare interventions. In addition, transport is limited[21], making it harder for people in these communities to reach trial sites. However, almost all sites had experience of remote methods of trial delivery, supporting development of a hybrid trial design to mitigate geographical accessibility challenges and increase participation opportunity. Sites reported challenges with workforce capacity, experience and expertise. We found that almost all UK regions had a tier 3 site with appropriate expertise and experience to support staff in less experienced sites. Provision of core funded staff to strategic tier 3 sites would provide visible and accessible local research mentorship[22] and training role-models[23] facilitating capacity growth. We found that tier 3 sites were more likely to be located in areas with higher ethnic diversity and deprivation. Strategic placement of core-funded staff within these sites would provide additional resource for engagement with these communities. Rater capacity limitations and known challenges with OFF state assessments[24], associated with limited overnight accommodation provision or ability to undertake home visit assessments, further supported development of a hybrid trial delivery model.

The EJS ACT-PD trial has an embedded digital measures sub-study. We found that approximately half of sites had experience of using digital technologies. Digital technologies can help enhance understanding of disease and deliverability of new therapies[25]; their use has risen exponentially due to the rise in remotely delivered trials following the COVID-19 pandemic[26]. Our survey identified issues with connectivity, software and NHS firewalls, similar to those previously reported[27]; the extent to which these issues impacts the use of digital measures in trials will need to be closely monitored.

There are several limitations to this study. Certain survey questions lacked granularity which may have hampered the identification of research gaps. For example, specialist imaging was identified as an area of concern in tier 1 sites, and although 20% of all sites indicated they could outsource access to the equipment from nearby acute trusts if needed, this was an open-ended question where sites could give feedback on any issues relating to imaging equipment. A direct question on outsourcing of equipment access may have provided more accurate data. Survey dissemination was in part dependent on distribution to sites identified via a national database of NIHR portfolio studies and utilised regional research delivery managers to encourage responses from known active research centres. This may have led to a bias towards more experienced PD sites and those active in nationally portfolio adopted research studies being represented in the dataset

## CONCLUSIONS

This work strongly advocates for the systematic scoping of research infrastructure and mapping of delivery capacity to population characteristics of interest can highlight training and delivery gaps and inform trial design and development of delivery strategies. We believe such approaches will result in more inclusive research and, ultimately, in more generalisable results.

## Supporting information

Data dictionary and coding definitions for quantitative data fields

Survey results by tier, challenges identified and the resulting mitigation strategies

EJS ACT-PD consortium members as of November 2024

## Data Availability

The datasets generated and analysed during the current study are not publicly available due the inclusion of NHS trust details but are available from the corresponding author on reasonable request.

## LIST OF ABBREVIATIONS

ACE III: Addenbrooke’s Cognitive Examination
ADL: Activities of Daily Living
BDI: Beck Depression Inventory
CRN: Clinical Research Network
CSF: Cerebral Spinal Fluid
DaTSCAN: Dopamine Active Transfer Scan
EJS ACT-PD: Edmond J Safra Accelerating Clinical Trials in Parkinson’s Disease
EQ-5D: EuroQol 5 Dimensions
IFWG: Infrastructure Working Group
IMD: Index of Multiple Deprivation
MAMS: Multi-arm multi-stage
MDS-UPDRS: Movement Disorder Society Unified Parkinson’s Disease Rating Scale
MMSE: Mini-mental state examination
MoCA: Montreal Cognitive Assessment
MRI: Magnetic Resonance Imaging
NHS: National Health Service
NIHR: National Institute for Health and Care Research
PD: Parkinson’s disease
PDQ-39: Parkinson’s Disease Questionnaire
PPIE: Patient and Public Involvement and Engagement
RDN: Research Delivery Network
REDCap: Research Electronic Data Capture
SCOPA-AUT: Scale for Outcomes in Parkinson’s disease for Autonomic symptoms
UK: United Kingdom

## DECLARATIONS

### Ethics approval and consent to participate

Not applicable

### Consent for publication

Not applicable

### Authors’ contributions

RP, M-LZ, SC, R E-D and GM created the survey on REDcap and facilitated the return of surveys from NHS Trusts. RP and M-LZ performed the analysis and categorisation of survey data. MB, LM and SW provided their personal expertise as lived experience experts. All authors comprised the EJS ACT-PD Infrastructure Working Group and developed site capability survey, the tiered site system and mitigations for infrastructure gaps identified in conjunction with the wider consortium.

## Acknowledgements

We would like to thank the wider EJS ACT-PD consortium (detailed in additional file 3) for their input into the survey. We would also like to thank all UK Research Delivery Managers for helping with survey dissemination, and each NHS site who took the time to complete the survey.

