## Supplementary material for "Scoping national research infrastructure to inform the design and delivery strategy for a platform clinical trial in Parkinson’s disease": Survey results by tier, challenges identified and the resulting mitigation strategies

Additional File 2: Survey results by tier, challenges identified and the resulting mitigation strategies

| Domain | Subdomain | Capabilities | Sites n(% of 97 survey responses received) | Tier 1 sites with capability n(% of 43 Tier 1 sites) | Tier 2 sites with capability n(% of 33 Tier 2 sites) | Tier 3 sites with capability n(% of 21 Tier 3 sites) | Delivery challenges identified | Mitigation strategy |
| --- | --- | --- | --- | --- | --- | --- | --- | --- |
| Trial experience | Trial experience | Interested in delivering PD trials in the future | 96(98.9) | 42(97.7) | 33(100.0) | 21(100.0) | None. Most sites have experience of delivering remote PD trials | Core funded staff to be located at strategic tier 3 sites to reach those from socially deprived and ethnic minority groups, and upskill and mentor less experienced sites |
|  |  | Experience with delivering remote trials | 89(91.8) | 38(88.4) | 30(90.9) | 21(100.0) |  |  |
|  |  | Capability to deliver neurological research | 84(86.6) | 31(72.1) | 32(97.0) | 21(100.0) |  |  |
|  |  | Experience in delivering PD clinical trials | 75(77.3) | 21(48.8) | 33(100.0) | 21(100.0) |  |  |
|  | Trial staff | Additional medical doctors to support trials (e.g. Research Fellows or Specialist Registrars) | 68(70.1) | 24(55.8) | 24(72.7) | 20(95.2) | There is a lack of available staff to support trial delivery across all tiers, particularly blinded staff in tier 1 and 2 sites |  |
|  |  | Dedicated PD research nurse or Allied Healthcare Professional | 52(53.6) | 12(27.9) | 23(69.7) | 17(81.0) |  |  |
|  |  | Sufficient staff to provide a blinded rater and backup | 44(45.4) | 10(23.3) | 19(57.6) | 15(71.4) |  |  |
| Rater expertise | PD scales | Modified Hoehn and Yahr | 66(68.0) | 13(30.2) | 32(97.0) | 21(100.0) | Sites are slightly more experienced in generic measures than PD specific ones. | Creation of a national rater training programme, in collaboration with the NIHR |
|  |  | Non-motor scales | 66(68.0) | 12(27.9) | 33(100.0) | 21(100.0) |  |  |
|  |  | MDS-UPDRS | 65(67.0) | 11(25.6) | 33(100.0) | 21(100.0) |  |  |
|  |  | PDQ-39 | 61(62.9) | 12(27.9) | 28(84.8) | 21(100.0) |  |  |
|  |  | OFF state assessments | 61(62.9) | 9(20.9) | 31(93.9) | 21(100.0) |  |  |
|  |  | Schwab and England ADL Scale | 57(58.8) | 15(34.9) | 24(72.7) | 18(85.7) |  |  |
|  |  | Scales for PD care partners | 52(53.6) | 7(16.3) | 28(84.8) | 17(81.0) |  |  |
|  |  | ON/OFF diaries | 46(47.4) | 6(14.0) | 20(60.6) | 20(95.2) |  |  |
|  |  | SCOPA-AUT | 41(42.3) | 4(9.3) | 21(63.6) | 16(76.2) |  |  |
|  | Non-PD scales | MoCA | 82(84.5) | 28(65.1) | 33(100.0) | 21(100.0) |  |  |
|  |  | MMSE | 81(83.5) | 28(65.1) | 32(97.0) | 21(100.0) |  |  |
|  |  | EQ-5D | 71(73.2) | 27(62.8) | 24(72.7) | 20(95.2) |  |  |
|  |  | Timed motor tests | 68(70.1) | 17(39.5) | 31(93.9) | 20(95.2) |  |  |
|  |  | ACE III | 67(69.1) | 21(48.8) | 26(78.8) | 20(95.2) |  |  |
|  |  | BDI | 59(60.8) | 16(37.2) | 25(75.8) | 18(85.7) |  |  |
|  |  | Smell tests | 23(23.7) | 2(4.7) | 8(24.2) | 13(61.9) |  |  |
|  | Digital outcomes | Wifi in area research is delivered | 95(97.9) | 42(97.7) | 33(100.0) | 20(95.2) | Tier 1 and 2 sites have limited experience of using in-person digital outcomes |  |
|  |  | Smartphone apps | 59(60.8) | 21(48.8) | 21(63.6) | 17(81.0) |  |  |
|  |  | Web-based assessments | 48(49.5) | 18(41.9) | 15(45.5) | 15(71.4) |  |  |
|  |  | Digital wearables | 46(47.4) | 13(30.2) | 16(48.5) | 17(81.0) |  |  |
|  |  | Video recorded outcomes for blinded rating | 27(27.8) | 3(7.0) | 14(42.4) | 10(47.6) |  |  |
| Trial facilities | Research space | Access to a clinical trials pharmacy | 88(90.7) | 34(79.1) | 33(100.0) | 21(100.0) | Most sites have pharmacy access but tier 1 and 2 sites are less likely to have dedicated research space | Utilise a hybrid model of in-person and remote study visits with the use of a central pharmacy to enable delivery even in tier 1 sites. No OFF state study visit requirements |
|  |  | Dedicated research rooms | 57(58.8) | 17(39.5) | 21(63.6) | 19(90.5) |  |  |
|  | Research accommodation | Ability to undertake home visits for research purposes | 56(57.7) | 28(65.1) | 17(51.5) | 11(52.4) | There is very little capacity for accommodation for research participants across all tiers |  |
|  |  | Nurse-observed overnight accommodation for research participants | 15(15.5) | 4(9.3) | 4(12.1) | 7(33.3) |  |  |
|  |  | Accommodation suitable for those with disabilities | 14(14.4) | 4(9.3) | 5(15.2) | 5(23.8) |  |  |
|  |  | Accommodation suitable for care partners of research participants | 10(10.3) | 4(9.3) | 3(9.1) | 3(14.3) |  |  |

Legend

Capability or experience in ≥75.1% of sites

Capability or experience in 50.1% - 75.0% of sites

Capability or experience in 25.1% - 50% of sites

Capability or experience in ≤25% of sites

Abbreviations: PD, Parkinson’s disease; MoCA, Montreal Cognitive Assessment; MMSE, Mini-mental state examination; EQ-5D, EuroQol 5 Dimensions; ADL, Activities of Daily Living; ACE III, Addenbrooke’s Cognitive Examination; BDI, Beck Depression Inventory; MDS-UPDRS, Movement Disorder Society Unified Parkinson’s Disease Rating Scale; PDQ-39, Parkinson’s Disease Questionnaire; SCOPA-AUT, Scale for Outcomes in Parkinson's disease for Autonomic symptoms
